# Women with breast implants have a higher rate of cardiac catheterization but a lower rate of percutaneous coronary intervention suggesting breast implant interference leads to higher false positive rates of cardiac stress imaging

**DOI:** 10.1101/2023.11.08.23298279

**Authors:** Mohammad Reza Movahed, Kyvan Irannejad, Emma Venard, Luke Keating, Mehrnoosh Hashemzadeh, Mehrtash Hashemzadeh

**Author notes:** Correspondent: M Reza Movahed, MD, PhD, FACP, FACC, FSCAI, Clinical Professor of Medicine, University of Arizona Sarver Heart Center, 1501 North Campbell Avenue, Tucson, AZ 85724.

## Abstract

**Background:** Breast implants cause attenuation during myocardial perfusion imaging (SPECT) and interfere with echocardiographic acoustic windows, which may cause false positive results. We hypothesized that false positive cardiac testing would lead to a lower rate of percutaneous intervention (PCI) in patients with breast implants who underwent cardiac catheterization.

**Methods:** Using ICD 10 codes for breast implants, cardiac catheterization, and percutaneous coronary interventions, we evaluated any association between these cardiac procedures and older adult women with breast implants.

**Results:** A total of 1,681,135 women 50 or older underwent cardiac catheterization and 792,760 of them underwent PCI. The total number of patients with breast implants who underwent cardiac catheterization was 1,980 (1.99%). The rate of cardiac catheterization was similar (2.99% vs 3.30%, p= 0.06). However, after multivariate adjustment, women with breast implants had a significantly higher rate of cardiac catheterization (OR: 1.12, CI: 1.00-1.25, P=0.05). Furthermore, women after undergoing cardiac catheterization had a lower unadjusted and adjusted rate of PCI (36.93% vs 47.08%, <0.001, unadjusted OR: 0.66, CI: 0.54-0.80, adjusted OR:0.79, CI: 0.64-0.97, p=0.02). Sensitivity analyses using different age cutoffs showed a similar pattern of findings.

**Conclusions:** Women with breast implants who underwent cardiac catheterization had significantly lower adjusted rates for PCI consistent with our hypothesis that breast implants may lead to higher false positive rates during cardiac ischemia testing. Therefore, women who consider breast implants should be informed about interference with cardiac diagnostic procedures.

## Introduction

Breast implants have become more popular recently. For example, one national study evaluating the prevalence of breast implants in the Netherlands based on chest radiograph data estimated the prevalence at approximately 3% (1). Silicone implants are approved by the FDA for cosmetic use (2–4). The most researched breast implants are silicone devices which have been used for decades (5). With advancing age, many women with breast implants require an increasing need for diagnostic cardiac testing for shortness of breath or chest pain. Studies employing social media data have highlighted high rates of concern about breast implant-related illness, including cardiac illness (6). However, interference of breast implants with cardiac diagnostic procedures is not much debated despite the increasing utilization of these [procedures in women with advancing age, Furthermore, a higher prevalence of atypical presentation of cardiac ischemia in women is well documented which would increase higher utilization of diagnostic cardiac testing(7). Commonly used methods for identifying hemodynamically significant coronary artery disease are radionuclide myocardial perfusion imaging (MPI), single photon emission computed tomography (SPECT), and positron emission tomography (PET) (8). The usage of echocardiography is another commonly performed diagnostic procedure in cardiology for the assessment of heart structures and function. Additionally, stress echocardiography is another frequently performed procedure for the evaluation of ischemia (9).

In women with large breasts can lead to significant breast attenuation thus contributing to a higher false positive rate in women with SPECT imaging (3,10). The presence of silicone or saline breast implants can dramatically increase the attenuation artifact during SPECT imaging leading to an increase in false positive rates in this population. (9,10). Furthermore, in the case of stress echocardiography utilization in this population for ischemia evaluation, breast implant causes severe acoustic shadow also limiting the diagnostic accuracy of stress echocardiography thus also increasing the false positive rate using stress echocardiography. (3,11). Despite recently increasing recognition of these challenges associated with diagnostic imaging in patients with breast implants, there has been limited empirical data evaluating rates of false positive outcomes in these patients. Therefore, the current study aims to evaluate rates of false positive cardiac testing for ischemia evaluation in patients with breast implants using the National Inpatients Sample (NIS) database. Specifically, we hypothesize that women with breast implants will have higher rates of cardiac catheterization but lower rates of percutaneous intervention (PCI), as an indicator of greater rates of false positive screening. To our knowledge, there is no prior study that shows the rate of PCI in patients with breast implants.

## Methods

For this study, the National Inpatient Sample (NIS) was utilized to conduct a retrospective analysis of archival data. The NIS is a collection of hospital inpatient databases from the Healthcare Cost and Utilization Project (HCUP). The purpose of the NIS was to create a set of databases from which national trends in healthcare utilization, quality of healthcare, and patient outcomes could be analyzed. All analyses were conducted following the implementation of population discharge weight. We evaluated elderly adult patients above the age of 50 for our study period between 2016-2020. From this database, patients with breast implants, cardiac catheterization, and percutaneous coronary interventions were identified via ICD 10 codes. We evaluated differences between women with and without breast implants in rates of cardiac catheterization and intervention. The NIS database can be found at www.hcup-us.ahrq.gov.

Furthermore, we performed multivariate analysis adjusting for age, race, diabetes, hypertension, hyperlipidemia, and smoking. Further secondary analysis was performed using different age cuts of points in adult women.

### Statistical analysis

Patient demographics, hospital, and clinical characteristics are reported in Tables 1 and 2. Primary analyses employed logistic regressions to compare women with breast implants to others in likelihood of binary clinical outcomes of cardiac catheterization and PCI. Odds ratios and 95% confidence intervals are calculated for continuous variables with 95% confidence intervals for categorical variables. Univariate linear regression for continuous variables was utilized.

**Table 1:** Adjusted and Unadjusted rates of Cardiac Catheterization By Age Cutoff.

|  | Unadjusted Model |  |  |  |  | Adjusted Models |  |  |  |
| --- | --- | --- | --- | --- | --- | --- | --- | --- | --- |
| Predictor Variable | Total | Breast Implant | Other | P-value | Odds Ratio (C.I) | Adjusted Odds Ratio (C.I) | P1-value | Adjusted Odds Ratio (C.I) | P2-value |
| **Age ≥ 18** | 1,871,335 | 2,250 | 1,869,085 |  |  |  |  |  |  |
| Age (Mean±SD) | 66.85±12.85 | 64.46±11.94 | 66.85±12.85 | <0.001 |  |  |  |  |  |
| Coronary Angiography | 2.20% | 1.99% | 2.20% | 0.04 | 0.90<br>(0.82-1.00) | 1.00 (0.90-1.10) | 0.92 | 1.31<br>(1.18-1.46) | <0.001 |
| **Age ≥ 30** | 1,861,065 | 2,235 | 1,858,830 |  |  |  |  |  |  |
| Age (Mean±SD) | 67±12.63 | 64.55±11.85 | 67.01±12.63 | <0.001 |  |  |  |  |  |
| Coronary Angiography | 2.61% | 2.11% | 2.61% | <0.001 | 0.80<br>(0.73-0.88) | 0.90<br>(0.81-0.99) | 0.026 | 1.18<br>(1.07-1.31) | 0.001 |
| **Age ≥ 40** | 1,822,885 | 2,175 | 1,820,710 |  |  |  |  |  |  |
| Age (Mean±SD) | 67.66±11.90 | 65.37±10.91 | 67.66±11.90 | <0.001 |  |  |  |  |  |
| Coronary Angiography | 3.14% | 2.58% | 3.14% | <0.001 | 0.82<br>(0.74-0.90) | 0.81<br>(0.73-0.89) | <0.001 | 1.08<br>(0.98-1.20) | 0.14 |
| **Age ≥ 50** | 1,681,135 | 1,980 | 1,679,155 |  |  |  |  |  |  |
| Age (Mean±SD) | 69.54±10.37 | 67.40±9.16 | 69.54±10.37 | <0.001 |  |  |  |  |  |
| Coronary Angiography | 3.30% | 2.99% | 3.30% | 0.06 | 0.90<br>(0.81-1.00) | 0.85<br>(0.76-0.94) | 0.002 | 1.12<br>(1.00-1.25) | 0.05 |

**Table 2:**
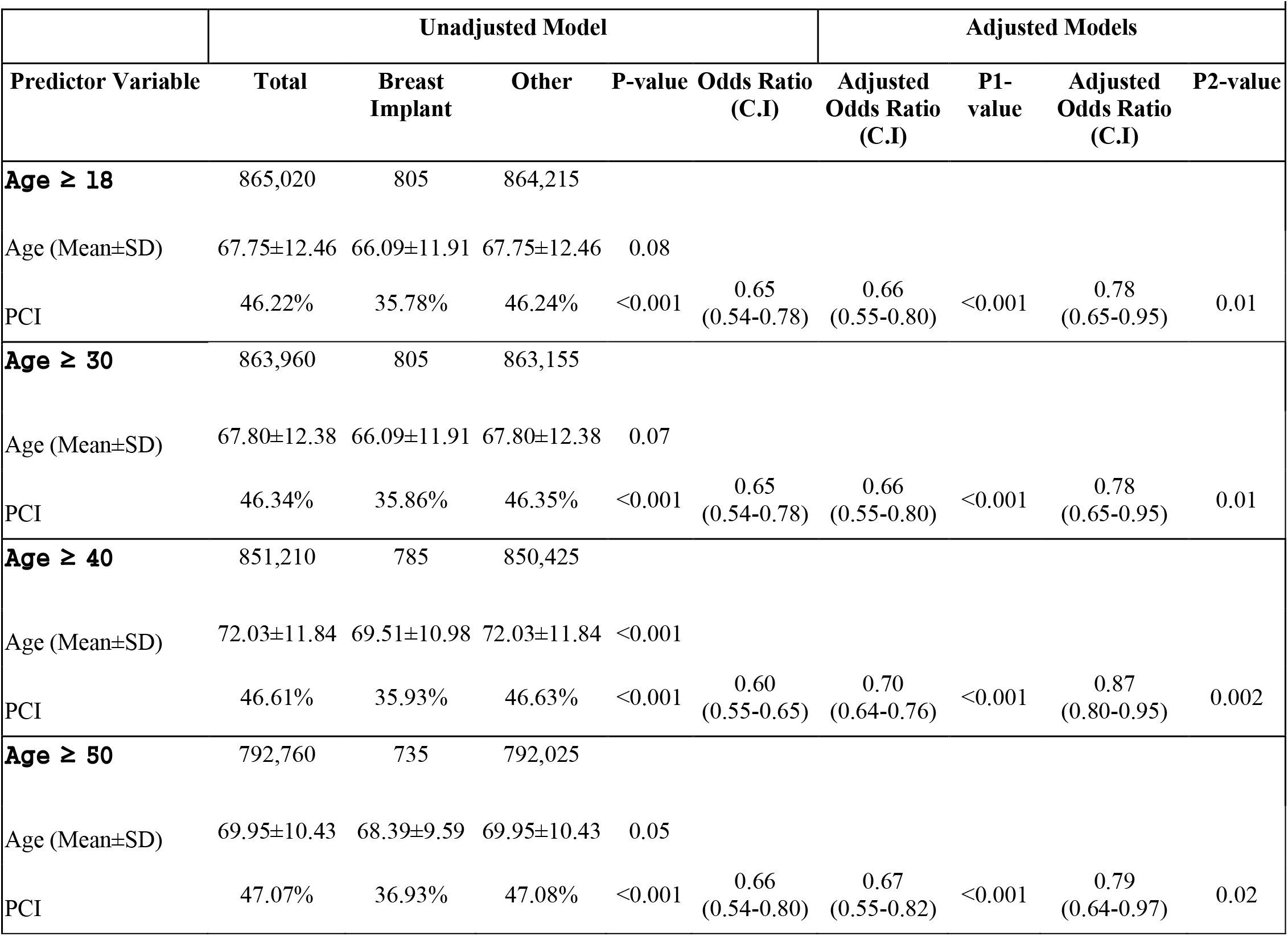
Adjusted and Unadjusted rates of Percutaneous Coronary Intervention (PCI) After Cardiac Catheterization By Age Cutoff.

Multivariate logistic regression ascertained the odds of binary clinical outcomes relative to patient procedures P-values are 2-sided and p<0.05 was considered statistically significant. Data were analyzed using STATA 17 (Stata Corporation, College Station, TX).

## Results

Primary inferential analyses were conducted on the sample of women using an age cutoff of 50 or older. This final analytic sample included a total of 1,681,135 women 50 or older who underwent cardiac catheterization, of whom 792,760 women 50 over older underwent, PCI identified from available ICD-10 codes. The total number of patients with breast implants who underwent cardiac catheterization was 1,980 (1.99%). Results of all analyses are presented in Tables 1 and 2. In unadjusted analyses, we found that women greater than 50 with breast implants did not significantly differ from others in the likelihood of cardiac catheterization (OR: 0.90, CI: 0.81-1.00, *p* = 0.06). However, after multivariate adjustment, women with breast implants had a significantly higher rate of cardiac catheterization (OR: 1.12, CI: 1.00-1.25, P=0.05). Furthermore, we found that women over 50 with breast implants who underwent cardiac catheterization were significantly less likely to have PCI in an unadjusted univariate analysis (OR: 0.66, CI: 0.54-0.80, *p* < .001). After adjusting for race, diabetes, hypertension, hyperlipidemia, STEMI, non-STEMI, and smoking status, women with breast implants had significantly lower adjusted PCI rates (adjusted OR: 0.79, CI: 0.64-0.97, p= 0.02).

Sensitivity analyses conducted using different age cutoffs showed a similar pattern of findings. In every subgroup age cut of points, we found that women with breast implants have higher adjusted rates of cardiac catheterization studying additional age cutoffs of 18, 30, and 40 or older (see Table 1 for results)[1]. Finally, our primary finding that women with breast implants have lower unadjusted, and adjusted rates of PCI following cardiac catheterization was found also using age cutoffs of 18, 30, and 40 or older (see Table 2 for results).

## Discussion

This study aimed to evaluate rates of false positive cardiac testing for ischemia evaluation in patients with breast implants using the National Inpatients Sample (NIS) database. To our knowledge, this is the first study to examine rates of PCI in patients with breast implants. We found that women with breast implants who underwent cardiac catheterization had significantly lower adjusted rates for PCI, consistent with our hypotheses. Results suggest that breast implants lead to higher false positive rates during cardiac ischemia testing. These findings were robust to age stratification, showing the same pattern of findings across selected age cutoffs.

Our findings are consistent with the limited available research studying interference from breast implants on cardiac ischemia testing. Several cases have reported that false-positive outcomes in gated SPECT examinations in women are frequently caused by breast attenuation artifacts (12). Case reports also show interference of breast implants with echocardiographic image acquisition and interpretation (3). It is unknown and unstudied what underlying physical characteristic of silicone breast implants interferes with the ultrasound beam during an echocardiogram. Silicone breast implants seem to block the penetration of ultrasonic rays, somewhat like air in the lung but to a smaller extent. Despite an increase in gain or a shift in the ultrasonic wave’s frequency, the low penetration seems to remain enduring and undetectable. Since silicone is less dense than calcium, it does not seem to significantly alter the shadowing that is visible in calcified formations(3). Three cases have been reported highlighting significant impairment of echocardiographic acoustic windows caused by breast implants (11). Further, the existing available data from the case series suggests that interference is not unique to silicone implants. For example, one case series reported on impaired myocardial SPECT imaging in two women with saline-containing implants as well as one woman with silicone implants (9). Taken together, there is support for the notion that breast implants lead to impaired SPECT imaging, but the specific mechanisms for impaired imaging and potential solutions remain unclear.

This study has implications for the reliability and validity of cardiac ischemia evaluation, which differs in patients with breast implants. Further, these findings can inform counseling of patients considering breast implants, as they may be at increased risk for false positive screening. Counseling and education in this context may be beneficial at the patient level in reducing unnecessary procedures and supporting diagnostic clarity (9). In addition, greater awareness of the impact of interference on the accuracy of cardiac ischemia testing may reduce healthcare costs for these patients. Therefore, women who consider breast implants should be informed about this important interference with their breast transplant that could lead to false positive screening and increasing utilization of invasive cardiac procedures thus increasing the risk of adverse events in this population.

## Conclusion

This is the first study examining false-positive screening for cardiac ischemia in patients with breast implants using the National Inpatients Sample (NIS). Further research is warranted to continue to examine sources of bias for cardiac screening in patients with breast implants. This study shows that women with breast implants who underwent cardiac catheterization have significantly lower adjusted rates of PCI. These findings are consistent with our hypothesis that breast implants may lead to higher false positive rates during cardiac ischemia testing. In addition, examination of the impact of education for patients about risks for interference associated with breast implants could inform best practices for counseling patients.

## Limitations

We cannot rule out other unknown causes of higher utilizations of cardiac catheterization in patients with breast implants warrant further investigations. We used ICD-10 coding to identify patients and their characteristics, which may be limited in accuracy.

## Data Availability

NIS data base publicly available

## References

1. de Boer M, van Middelkoop M, Hauptmann M, van der Bijl N, Bosmans JAW, Hendriks-Brouwer N, et al. Breast implant prevalence in the dutch female population assessed by chest radiographs. Aesthet Surg J. 2020 Jan 29;40(2):156–64.

2. Cook RR, Delongchamp RR, Woodbury M, Perkins LL, Harrison MC. The prevalence of women with breast implants in the United States--1989. J Clin Epidemiol. 1995 Apr;48(4):519–25.

3. Movahed M-R. Interference of breast implants with echocardiographic image acquisition and interpretation. Cardiovasc Ultrasound. 2007 Feb 23;5:9.

4. Le GM, O’Malley CD, Glaser SL, Lynch CF, Stanford JL, Keegan TH, et al. Breast implants following mastectomy in women with early-stage breast cancer: prevalence and impact on survival. Breast Cancer Res. 2005;7(2):R184–93.

5. Hedén P, Nava MB, van Tetering JPB, Magalon G, Fourie LR, Brenner RJ, et al. Prevalence of rupture in inamed silicone breast implants. Plast Reconstr Surg. 2006 Aug;118(2):303–8; discussion 309.

6. Tang SYQ, Israel JS, Afifi AM. Breast implant illness: symptoms, patient concerns, and the power of social media. Plast Reconstr Surg. 2017 Nov;140(5):765e–6e.

7. Azis KA, Al-Chalabi MMM, Mat Johar SFN, Wan Sulaiman WA. Atypical chest pain in a patient with breast implant. Cureus. 2023 Apr 18;15(4):e37751.

8. Al Badarin FJ, Malhotra S. Diagnosis and Prognosis of Coronary Artery Disease with SPECT and PET. Curr Cardiol Rep. 2019 May 18;21(7):57.

9. Stinis CT, Lizotte PE, Movahed M-R. Impaired myocardial SPECT imaging secondary to silicon- and saline-containing breast implants. Int J Cardiovasc Imaging. 2006 Mar 15;22(3–4):449–55.

10. Mieres JH, Shaw LJ, Arai A, Budoff MJ, Flamm SD, Hundley WG, et al. Role of noninvasive testing in the clinical evaluation of women with suspected coronary artery disease: Consensus statement from the Cardiac Imaging Committee, Council on Clinical Cardiology, and the Cardiovascular Imaging and Intervention Committee, Council on Cardiovascular Radiology and Intervention, American Heart Association. Circulation. 2005 Feb 8;111(5):682–96.

11. Movahed M-R. Impairment of echocardiographic acoustic window caused by breast implants. Eur J Echocardiogr. 2008 Mar;9(2):296–7.

12. Movahed M-R. Attenuation artifact during myocardial SPECT imaging secondary to saline and silicone breast implants. Am Heart Hosp J. 2007;5(3):195–6.

